# Fast and Accurate Maximum-Likelihood Estimation of Multi-Type Birth-Death Epidemiological Models from Phylogenetic Trees

**DOI:** 10.1101/2022.08.02.22278328

**Authors:** Anna Zhukova, Frédéric Hecht, Yvon Maday, Olivier Gascuel

**Author notes:** Corresponding authors (AZ), (OG).

## Abstract

Multi-type birth-death (MTBD) models are phylodynamic analogies of compartmental models in classical epidemiology. They serve to infer such epidemiological parameters as the average number of secondary infections *R*_*e*_ and the infectious time from a phylogenetic tree (a genealogy of pathogen sequences). The representatives of this model family focus on various aspects of pathogen epidemics. For instance, the birth-death exposed-infectious (BDEI) model describes the transmission of pathogens featuring an incubation period (when there is a delay between the moment of infection and becoming infectious, as for Ebola and SARS-CoV-2), and permits its estimation along with other parameters.

With constantly growing sequencing data, MTBD models should be extremely useful for unravelling information on pathogen epidemics. However, existing implementations of these models in a phylodynamic framework have not yet caught up with the sequencing speed. Computing time and numerical instability issues limit their applicability to medium data sets (*≤* 500 samples), while the accuracy of estimations should increase with more data.

We propose a new highly parallelizable formulation of ordinary differential equations for MTBD models. We also extend them to forests to represent situations when a (sub-)epidemic started from several cases (e.g., multiple introductions to a country). We implemented it for the BDEI model in a maximum likelihood framework using a combination of numerical analysis methods for efficient equation resolution. Our implementation estimates epidemiological parameter values and their confidence intervals in two minutes on a phylogenetic tree of 10 000 samples. Comparison to the existing implementations on simulated data shows that it is not only much faster, but also more accurate. An application of our tool to the 2014 Ebola epidemic in Sierra-Leone is also convincing, with very fast calculation and precise estimates. As MTBD models are closely related to Cladogenetic State Speciation and Extinction (ClaSSE)-like models, our findings could also be easily transferred to the macroevolution domain.

---

The interaction of epidemiological and evolutionary processes leaves a footprint in pathogen genomes. Phylodynamics leverages this footprint to estimate epidemiological parameters, such as the average number of secondary infections, *R*_*e*_ (Grenfell et al., 2004; Volz et al., 2013). It relies on models that bridge the gap between traditional epidemiology and sequence data. Under these models, the parameter inference is drawn from topology and branch lengths of pathogen phylogenetic trees (i.e., genealogies of the pathogen population, approximating the transmission trees) combined with metadata on the samples. This is particularly useful for emerging epidemics, for which not enough data (e.g., incidence curves) might yet be gathered for accurate estimations with classical epidemiological methods. Rapidly growing genetic data coupled with phylodynamic estimations can provide valuable insights at an early stage of the epidemic spread and help prevent it (e.g., accurate estimation of *R*_*e*_ is crucial for adjusting potential non-pharmaceutical interventions, such as lockdowns).

Phylodynamic models can be classified into two main families: coalescent (Volz et al., 2009; Drummond et al., 2005; Pybus et al., 2000) and birth-death (BD) (Kendall, 1948; Maddison et al., 2007; Stadler, 2009, 2010). Coalescent models are often preferred for estimating deterministic population dynamics, however BD models are better adapted for highly stochastic processes, such as the dynamics of emerging pathogens (Macpherson et al., 2021). In BD models, births represent pathogen transmission events, while deaths correspond to becoming non-infectious (e.g., due to healing, self-isolation, starting a treatment, or death). Models of the BD family are phylodynamic analogies of compartmental models in classical epidemiology (e.g., SIR, Susceptible-Infectious-Recovered (Hethcote, 2000)). Many extensions of the classical BD model with incomplete sampling (BDS (Stadler, 2009)) were developed over time, including multi-type birth-death (MTBD) models (Stadler and Bonhoeffer, 2013). They add a population structure to the classical birth-death process by allowing for different types of individuals. A particularly useful representative of the MTBD family is the birth-death exposed-infectious (BDEI) model (Stadler et al., 2014). It was designed for pathogens featuring an incubation period between the moments of infection and of becoming infectious, e.g., Ebola and SARS-CoV-2. It is closely related to the SEIR (Susceptible-Exposed-Infectious-Recovered (Hethcote, 2000)) model, widely used in classical epidemiology.

In MTBD framework, the evolution of a transmission tree is described with a system of master differential equations with respect to global time. The model parameters can be estimated with maximum-likelihood (Stadler and Bonhoeffer, 2013) or Bayesian methods (Bouckaert et al., 2019) by exploring the likelihood (or posterior probability) landscape of trees. However, the closed form solution of the master equations exists only for the initial BDS model (Stadler, 2009), while for its extensions (like the BDEI model and MTBD models in general) the master equations for likelihood calculation need to be resolved with numerical methods. The complexity of the master equations and their boundary conditions (which recursively depend on the tree evolution later in time), make their numerical resolution challenging and time consuming (Scire et al., 2022; Voznica et al., 2022). The trade-off between the complexity of the biological questions a model can address and the computational speed for its parameter estimation is crucial in phylodynamics. On one hand, denser sampling should improve the accuracy of parameter estimations with complex models. On the other hand, denser sampling leads to larger data sets (thousands of samples), while computational issues often limit model applicability to medium or small ones (hundreds of samples). Calculations become time-consuming and numerically challenging (e.g., due to underflow issues) as tree size increases, resulting in numerical instability and inaccuracy (Scire et al., 2022; Voznica et al., 2022). Existing likelihood-based implementations of MTBD models (Stadler and Bonhoeffer, 2013; Bouckaert et al., 2019; Scire et al., 2022) can handle trees of medium size. In (Voznica et al., 2022) we proposed PhyloDeep, a likelihood-free deep-learning-based solution to the numerical instability issue. While being very efficient and accurate at the prediction stage, this approach however requires a computationally heavy training stage: Millions of trees covering a wide parameter range (where the real data is expected to fall) need to be simulated for training the deep learning predictor.

In the macroevolution domain, there exist several models that are closely related to the MTBD models. These are the models of the State Speciation and Extinction (SSE) family, in which the births correspond to species specifications and the deaths correspond to extinctions. The main difference is that in epidemiological models sampling happens though time, while in the macroevolution ones it usually occurs at present (at the extant species). Important representatives of SSE model family include the Binary SSE (BiSSE (Maddison et al., 2007)) model, which introduced two compartments with a possibility of anagenetic state change between them (i.e., along the tree branches), its extension to any number of states (multiple SSE, MuSSE (FitzJohn et al., 2009)), and the cladogenetic SSE (ClaSSE (Goldberg and Igić, 2012)) model, which introduced a possibility of cladogenetic state changes (i.e., when one of the offsprings may have a different state from its parent’s one right after the speciation event). In a recent work Louca and Pennell (2020) described a general mathematical framework for efficient likelihood calculation of these types of models, based on the “flow”, and implemented it for the MuSSE-like models but not for the ClaSSE-like ones. However, macroevolutionary analogues of the BDEI and general MTBD models belong to the ClaSSE-like family (as they permit a donor and a recipient to be in different states at the moment of transmission), and an efficient parameter estimator for these models on very large trees is currently lacking.

In this study we introduce a likelihood-based approach that intends to improve the accuracy and reduce the likelihood computation time of MTBD models. We propose a new formulation of the MTBD master equations that (i) removes the recursive dependency between child and parent nodes in the tree, hence permitting their parallel computation, and (ii) avoids numerical issues that could arise from very small boundary condition values. Under our approach, the master equations are resolved in parallel for different tree nodes, and then combined into the tree likelihood. In the general MTBD case the combination step is performed in a computationally light recursive way. However, we identified a subclass of MTBD models (including the BDEI model) whose likelihood formulae can be expressed in a non-recursive manner, thus allowing for even simpler calculations. Additionally, we extend the MTBD models applicability from single trees to forests. Forests could correspond to multiple introductions of the epidemic to the region of interest, or to a health policy change, which led to a new epidemic stage starting with several cases.

We applied our findings to the BDEI model and implemented its parameter estimator PyBDEI, employing targeted numerical analysis methods for accurate and fast resolution of its equations. We show the accuracy and speed of PyBDEI on simulated data and compare it to the gold standard Bayesian tool BEAST2 (Bouckaert et al., 2019) and the deep-learning-based tool PhyloDeep (Voznica et al., 2022). We find that our approach outperforms the competitors and makes the BDEI model applicable to very large data sets. Lastly, we apply PyBDEI to infer the epidemiological parameters that shaped the Ebola epidemic in Sierra-Leone in 2014. Our estimator is freely available at github.com/evolbioinfo/bdei.

## 1 MTBD models and their special case, the BDEI model

In a pathogen transmission tree *𝒥* (approximated by a time-scaled pathogen phylogeny) the tips represent sampled pathogens, patient state transitions occur along the branches, and bifurcations (i.e., internal nodes) correspond to transmissions (Fig. 1). The tree branch lengths are measured in units of time, where *T* is the time that passed between the beginning of the (sub-)epidemic (*t*_*start*_, corresponding to the time of the root in Fig. 1) and the last sampled tip.

**Figure 1.**
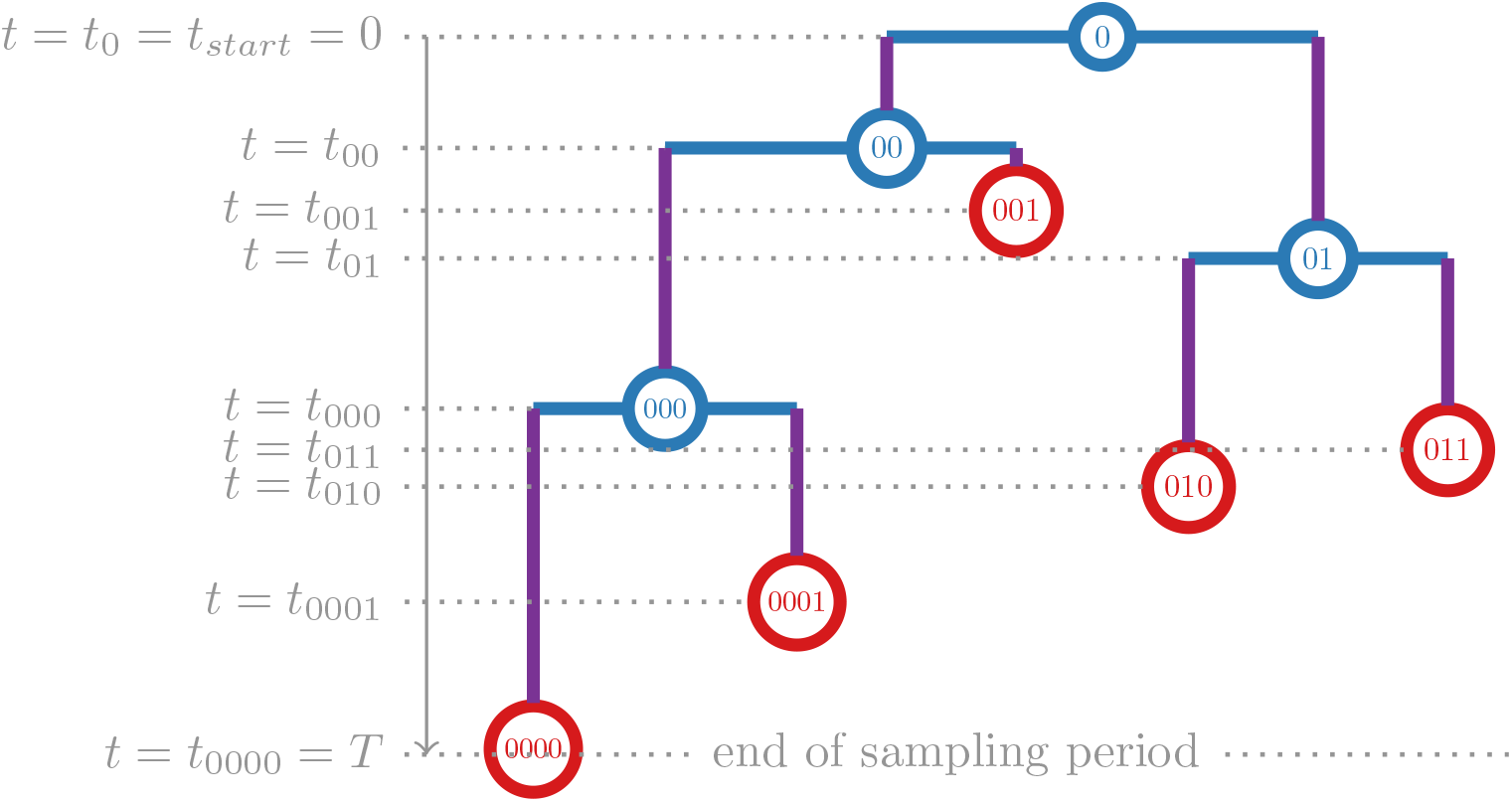
A transmission tree *𝒥* with *n* = 5 external nodes (i.e., tips, which correspond to sampling events: 0000, 0001, 001, 010, 011), *n* − 1 = 4 internal nodes (which correspond to transmissions: 0 (the root) and 00, 01, 000) and 2*n* − 2 = 8 branches (plus the root branch of zero length). Time *t* starts at the beginning of the (sub-)epidemic (here represented by the root of the tree, *t* = 0) and goes till the last sampled tip. The times of the nodes are shown on the left, e.g., *t*_0001_ is the time of tip 0001 (when 0001’s pathogen was sampled). *T* corresponds to the end of the sampling period (when the most recent tip, 0000, was sampled).

**The basic BDS model** (Stadler, 2009) has only one state: *infectious I*. An individual in state *I* can transmit their pathogen to another individual (whose state will be also *I*) at a constant average rate *λ*, or stop being infectious at a constant average rate *ψ* (due to treatment start, healing, self-isolation or death). After stopping being infectious, the individual and their pathogen exit the study, at which point the pathogen might get sampled with a probability *ρ*. The sampling is incomplete: an infectious individual may be removed from the system without being sampled (i.e., unobserved in the transmission tree), for example due to healing. The BDS model permits inference of such important epidemiological parameters as:

- *effective reproduction number* 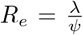, expected number of individuals directly infected by an infectious case;
- *infectious time* 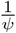, time during which an infectious individual can further spread the epidemic.

The BDS model is asymptotically unidentifiable (see Remark 3.4 in (Stadler, 2009)), but to become identifiable it requires one of the parameters to be fixed.

**MTBD models** (Stadler and Bonhoeffer, 2013) add population structure by allowing different individual states, transmissions between them and state changes. A general MTBD model with *d* individual states has 2*d*^2^ + *d* parameters: An individual in state *k* (*k ∈ {*1, …, *d}*) can be removed at a constant average rate *ψ*_*k*_ (with pathogen sampling probability *ρ*_*k*_), change their state to state *l* (*l ∈ {*1, …, *d}*) at a constant average rate *μ*_*kl*_ (where *μ*_*kk*_ = 0), and transmit their pathogen to an individual in state *l* at a constant average rate *λ*_*kl*_. The time between events of the same type is hence modelled with exponential distribution.

**The BDEI model** (Stadler et al., 2014) (Fig. 2), for example, is a special case of MTBD models that adds a second possible state to the state *I*: *exposed E*, an individual who is already infected but not yet infectious (cannot transmit), and will eventually become infectious.

**Figure 2.**
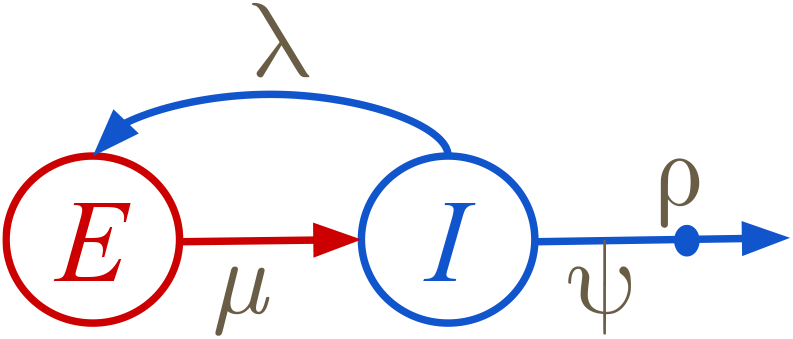
The BDEI model. An individual in exposed state *E* becomes infectious at a rate *μ*. An infectious individual *I* transmits the pathogen at a rate *λ* (hence creating a new exposed individual *E*), and gets removed at a rate *ψ* (decreasing the number of infectious individuals *I*). Upon removal, the individual’s pathogen might be observed with a probability *ρ*. Note, that the BDEI model does not include a susceptible state *S* (as for example SEIR) and makes the assumption that the susceptible population is unlimited (as for example in the beginning of an epidemic, or when the removed individuals could get reinfected).

Under the BDEI model, the only allowed transmissions are from *I* to *E*: At the moment of a transmission, the transmitter is always in state *I*, while the recipient is in state *E*. Hence the only non-zero transmission rate is *λ*_*IE*_ = *λ*, while the other 3 transmission rates are trivial: *λ*_*II*_ = *λ*_*EI*_ = *λ*_*EE*_ = 0. As we typically do not have the information to distinguish a transmitter from a recipient in a phylogenetic tree (which approximates the transmission tree), we have to consider both possibilities during parameter estimation. For the MTBD models where multiple states can transmit, the number of possibilities increases combinatorially.

Under the BDEI model, we assume that only the individuals in state *I* can exit the study (at rate *ψ*_*I*_ = *ψ*) and be detected and sampled (with a probability *ρ*_*I*_ = *ρ*): For instance, for many pathogens with an incubation period, the detection is triggered by the onset of symptoms, which in turn happens in the infectious state. Hence all the BDEI tree tips are in state *I*, and *ψ*_*E*_ = *ρ*_*E*_ = 0.

In addition to *R*_*e*_ and infectious time, the BDEI model permits inference of a third epidemiological parameter:

- *incubation period* 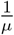, time between the infection and becoming infectious.

The incubation period can be expressed via becoming infectious rate *μ*_*EI*_ = *μ*, corresponding to a state transition from *E* to *I*. The inverse state change is not allowed: *μ*_*IE*_ = 0.

MTBD models, as extensions of the BDS model, are asymptotically unidentifiable and require one of their parameters to be fixed in order to become identifiable. In practice, it is often the sampling probability, as it may be approximated from epidemiological data (e.g., the proportion of sampled cases among the declared ones) or the infectious time (estimated from observations of infected cases).

### 1.1 Master equations

In the standard MTBD master equations proposed by Stadler and Bonhoeffer (2013), time goes backward from the last sampling event (the most recent tip in the tree) till the beginning of the epidemic. These equations permit calculation of the likelihood density of the data (observed tree) given the model parameter values Θ. In the general MTBD case, the observed tree, reconstructed from sampled pathogen genomes, differs from the real transmission tree: the states of its internal nodes (corresponding to transmissions) are unknown, we cannot distinguish between the transmitter and the recipient branches, the moments of state changes are also unknown, and due to incomplete sampling some parts of the real transmission tree are unobserved in the reconstructed tree. We therefore need to integrate over all possibilities while calculating the likelihood. The BDEI model is a slightly simpler case as only infectious individuals can transmit or get sampled, hence all the node states are known (*I*). For the BDEI model Θ = {*μ, λ, ψ, ρ*}.

In System (1) we show the MTBD master equations for a model with *d* states (for the BDEI model *d* = 2), however presenting them with the time *t* going forward from the time of the epidemic start (*t* = *t*_*start*_ = 0, i.e., tree root) to the time of the last sampled tip (*t* = *T*). These equations describe the likelihood density functions (LDFs) 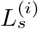 (*t*) of evolving as in the reconstructed tree, starting at time *t* in state *s* (for the BDEI model *s ∈ {E, I}*) on a branch connecting a node *i* to its parent, and till the end of the sampling period. The boundary condition is defined at time *t* = *t*_*i*_ (i.e., at the node *i*). To account for incomplete sampling, the system also includes the probabilities *U*_*s*_(*t*) of evolving unobserved till the time *T*, starting at time *t* in state *s*.

From now on, we use the following notation: the id of the root node is 0; the ids of its children are 00 and 01; and, by extension, the children of a node *i* (if they exist) have ids *i*0 and *i*1 (as in Fig. 1). Additionally if the state of a node *i* is known, then we will name it *si*. (For example for the BDEI model *si* = *I* ∀*i*.)

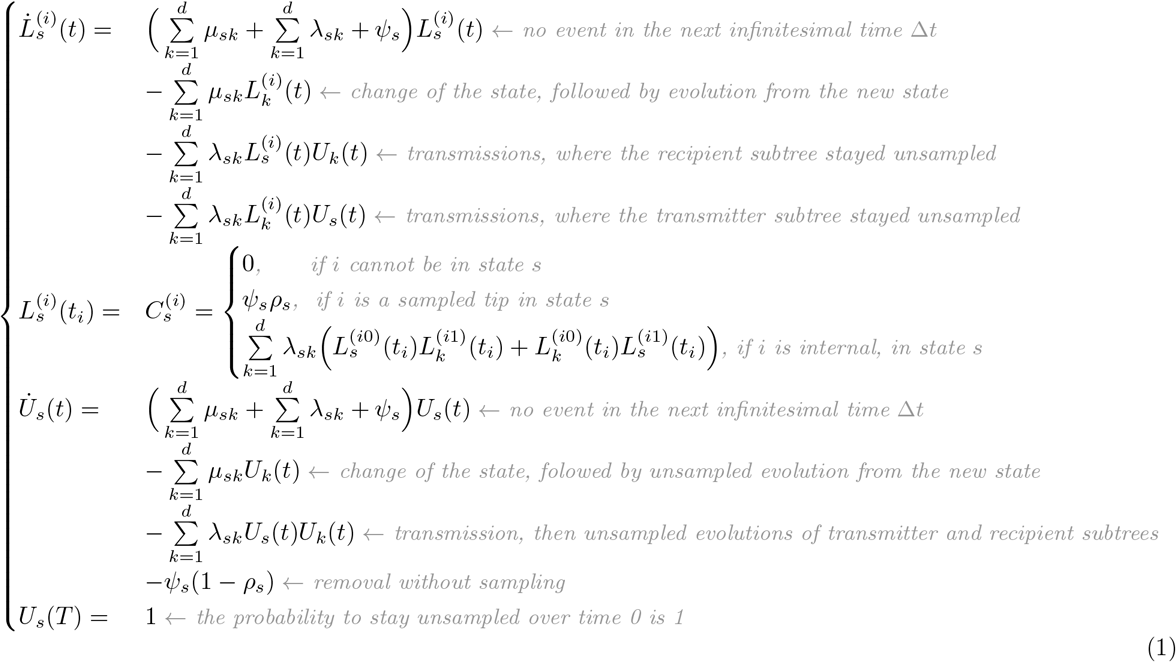

### 1.2 Tree likelihood density

The likelihood density of a tree *𝒥* for given parameter values Θ is then calculated as the LDF at time *t* = *t*_0_ = 0 on the root, whose id is 0 and whose state is *s*0:

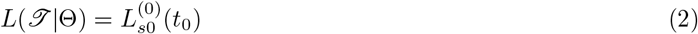

So far, we assumed that the epidemic started directly with the first transmission, however we can relax this assumption. The root of the tree in Figure 1 is placed at *t* = 0, and does not have a branch (its length is zero). Allowing for a non-zero root branch corresponds to an epidemic start some time before the first transmission (*t*_*start*_ = 0 *< t*_0_). This implies that the state of the individual represented by the root branch at time *t* = 0 is unknown, and all possible states should be considered (Eq. (3)). The same formula applies to cases where the root state is unknown. Assuming that the relative number of individuals in each state is at equilibrium, we can calculate the weight *π*_*s*_ of each possible state *s* (derived in (Stadler et al., 2013) for the general MTBD case and in Equation (16) for the BDEI model).

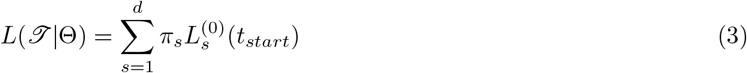

Root’s LDF *recursively* depends on the LDFs of the child node branches via the boundary condition 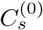 (System 1), and hence is calculated with a pruning algorithm (Felsenstein, 1973) while climbing the tree from tips till the root. Therefore when parallelized to maximum, it still requires *O*(*h*_*𝒥*_) consecutive steps, where *h*_*𝒥*_ stands for the height of the tree *𝒥* and depends on its topology: (balanced tree) *log*(*n*) *≤ h*_*𝒥*_ *≤ n* (ladder-like tree). At each step System (1) needs to be resolved for the corresponding nodes. Moreover, the values of 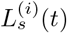 at internal nodes and their boundary conditions 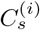 progressively become smaller as getting deeper in the tree, due to successive additions and multiplications of the LDF values. In trees with many tips, this might lead to numerical underflow, and hence such measures as rescaling need to be taken (Berger and Stamatakis, 2009; Defour, 2010; Scire et al., 2022).

### 1.3 Extension to forests

In some cases, the assumption that a (sub-)epidemic started with one infected individual might be too constraining. For instance, there could be multiple pathogen introductions to a country of interest (e.g., while in China the SARS-CoV-2 epidemic is commonly assumed to have started with one case, there were multiple independent introductions to other countries (Zhukova et al., 2020)). This scenario is depicted in Figure 3b.

**Figure 3.**
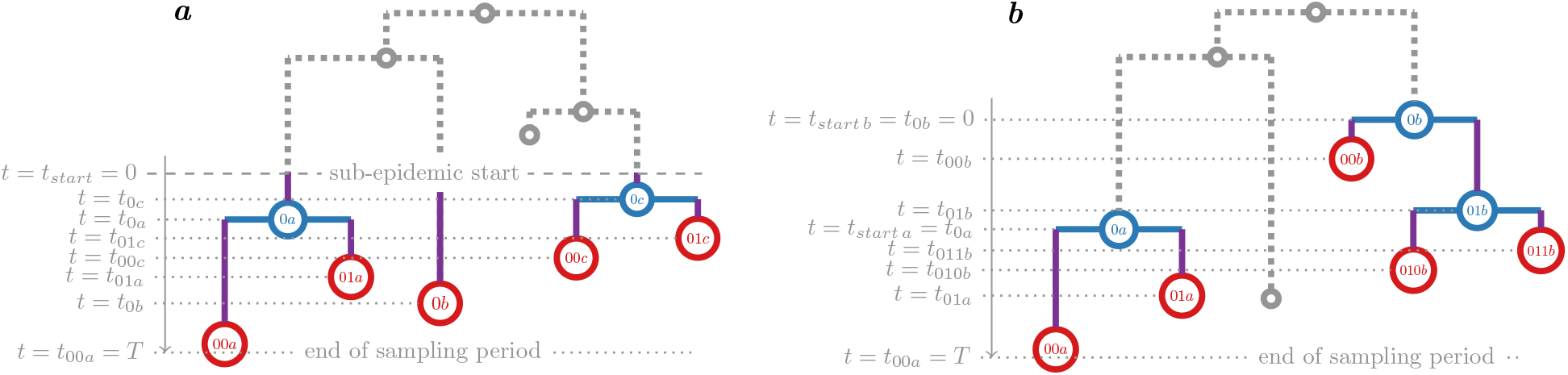
Forest *ℱ* representing a (sub-)epidemic that started with multiple infected cases. **(a)** The observed forest trees (*a, b* and *c*), corresponding to three different initial infected cases, are shown in color. All the forest trees start at the same time *t* = *t*_*start a*_ = *t*_*start b*_ = *t*_*start c*_ = *t*_*start*_ = 0). This scenario can correspond to a change of health policies leading to a change in parameter values (e.g., sampling). Such a change corresponds to a new stage of the epidemic, starting from several infected cases from the previous stage (shown in dashed gray). **(b)** The observed forest trees (*a, b*), corresponding to two different initial infected cases, are shown in color. They start at different times (*t*_*start b*_ *< t*_*start a*_). This scenario can correspond to multiple introduction to the same country from other countries (shown in dashed gray).

Another example is a change of health policies leading to a change in parameter values (e.g., sampling). Such a change corresponds to a new stage of the epidemic, starting from several infected cases from the previous stage. This scenario is depicted in Figure 3a. In Bayesian settings, the situations when the system behaviour (and parameters) change over time, are modeled via skyline methods. Stadler et al. (2013) developed the one-state Bayesian birth-death skyline plot that divides the time into intervals and allows for different piece-wise constant rates on them. Kühnert et al. (2016) combined the MTBD model with the skyline to allow for both piece-wise constant rate changes over time and multiple individual types. The skyline approach therefore relies on a single tree, but estimates a separate set of parameters for each time interval, all under the same model. As the number of parameters increases with multiple skyline intervals, MTBD-skyline models therefore require more data and computational time for their accurate estimation, and are more prone to numerical instability than the classical MTBD models.

We propose a simpler alternative, where the (sub-)epidemic starts with multiple individuals (not necessarily at the same time) and leads to a forest *ℱ* of *f* observed trees: *𝒥*_1_, …, *𝒥*_*f*_. The forest might also include a certain number *u* of unobserved trees, i.e., individuals who were infected at the beginning of the (sub-)epidemic, but whose trees stayed unobserved as none of their tips got sampled. This can be incorporated in the likelihood calculation. Forest likelihood formula hence combines the likelihoods of *f* observed and *u* hidden trees, and can be represented in logarithmic form (4). Tree likelihood formula is its special case, where *f* = 1 and *u* = 0.

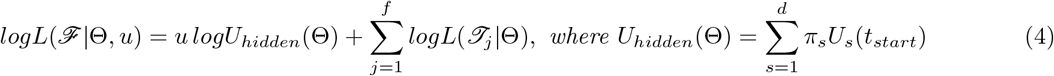

In Equation (4) we assumed that all the *f* +*u* sub-epidemics in the forest *ℱ* started at the same time (*t*_*start j*_ = *t*_*start*_ = 0 ∀*j ∈* {1, …, *f*}). This condition can be easily relaxed by replacing zeros with the corresponding tree starting times for observed and unobserved tree evolutions. As in practice we do not know the starting times of the unobserved trees if the sub-epidemics could start at different times, we approximate the unobserved tree starting times with the mean of the observed tree starting times: 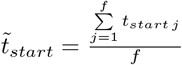 :

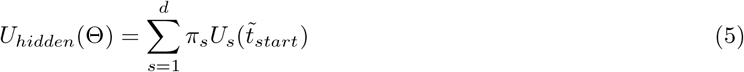

In our parameter estimator implementation for the BDEI model (see “Efficient parameter and CI estimation for the BDEI model” section), mean can be replaced with median, maximum or minimum of the observed tree times, via a user-specified parameter *u _policy*.

For given model parameter values Θ we can estimate the number of hidden trees *u* from the number of observed trees *f* as:

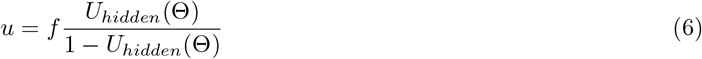

Hence, working with forests does not add an additional parameter to likelihood estimation. Moreover, as our simulations show (see “Performance on simulated data and comparison to other tools” section), the value of *u* has little impact on the parameter estimation. However, in some cases (e.g., change in health policy and thus of parameter values), it might be better to estimate u based on external data (e.g., number of cases at *t* = *t*_*start*_), rather than assuming that the parameter values predating the trees in the forest were the same as those in the forest. We explore both approaches in the “Application: Ebola in Sierra-Leone” section.

Using forests permits estimation of the model parameters on the last skyline interval without the restriction that the epidemic followed the same model before this interval (i.e., the top part of the tree, which includes the common ancestors of the forest roots, Fig. 3a). It reduces the number of parameters to those of the last interval. It also permits estimation of parameters for a (sub-)epidemic that started with several individuals but not at the same time (e.g., due to multiple introductions to a country, Fig. 3b).

## 2 Avoiding numerical problems and parallelizing calculations

In this section we introduce a way to rewrite LDFs in System (1) that permits (i) obtaining simpler boundary conditions to avoid potential numerical issues during resolution of equations; and (ii) removing recursion and resolving equations for each tree node in parallel, hence speeding up the calculations.

System (1) has several properties. First of all, its subsystem that defines unobserved probabilities (*U*_1_(*t*), …, *U*_*d*_(*t*)) is self-defined, and hence can be calculated independently from the rest. Secondly, in the subsystem that defines observed LDFs 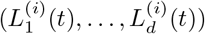 the right-hand side of the differential equations is a sum where each element is linear with respect to one of the 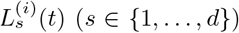, and this sum does not contain any free term. This condition implies that if we rescale 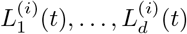 by a common factor, the differential equations will not change. Moreover, if the boundary conditions for all states but one are zero 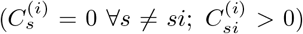, the rescaling will change the boundary condition only for 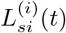. The latter is the case when tree node states are known, for example for the BDEI model, under which all nodes are in state *I*.

Assuming the state of the node *i* is known (*si*), let us define 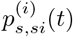 as:

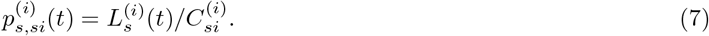

Then the differential equations for 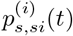 will only differ from those for 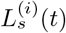 in the boundary condition for *s* = *si*, which is 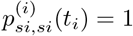 (the other boundary conditions stay zero). Conceptually, 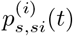 is a probability of an individual evolving as on an observed branch that connects a node *i* to its parent, starting at time *t* in state *s* on this branch and finishing at time *t*_*i*_ in state *si* (without taking into account *i*’s subtree and the event at node *i*).

Solving the master equations for 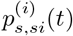 instead of 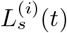 permits us to both (i) remove the recursive dependency between child and parent nodes (during ODE resolutions); and (ii) avoid numerical issues that could arise from very small values of the boundary condition of 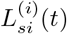, which is particularly pertinent for large trees. The calculation of 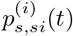 can be done in parallel for each node *i*. Hence, when parallelized to maximum, the number of parallel master equation resolutions becomes constant. In the general case, where the tree node states are unknown, all the possibilities (*si* = *k* ∀*k* ∈ {1, …, *d*}) for each node *i* can be considered separately and in parallel. Calculating 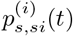 for all the *d ×d* possible combinations of (*s, si*) corresponds to the flow matrix G calculation in the general formulation recently proposed by Louca and Pennell (2020).

In the case where all node states are known (e.g., from metadata or due to model itself, as for the BDEI case), using 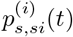 instead of 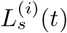 also permits us to express tree likelihood for model parameters Θ in a non-recursive way, and easily transform it to a logarithmic form (8) (to avoid underflow issues while multiplying small numbers at the likelihood combination step). In Materials and Methods we show its equivalence to the recursive representation (2).

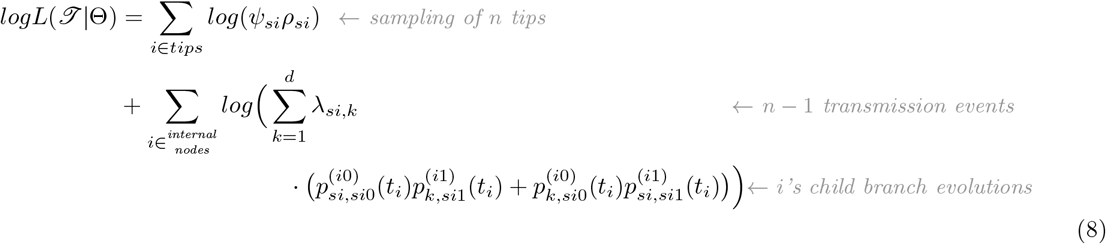

For the general case, the combination of different internal node state configurations into tree likelihood formula need to be performed with a pruning algorithm (Felsenstein, 1973). The likelihood-combining tree traversal starts from the tips and climbs the tree till the root, while calculating a subtree LDF 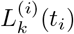 for each visited node *i* for each possible state *k*:

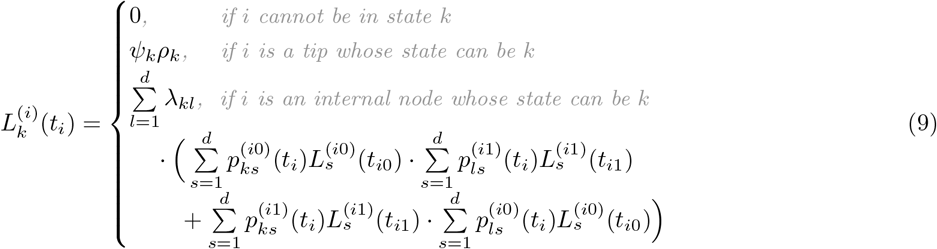

Note that unlike the known-tree-node-state likelihood (Eq. (8)), the recursive unknown-tree-node-state likelihood (Eq. (9)) does not allow for an easy logarithmic representation, and hence is prone to underflow issues. Its calculation on large trees therefore requires additional small number rescaling techniques as recently described in (Scire et al., 2022) and common in phylogenetic inference. However, unlike in the original MTBD representation, master equation resolutions (for 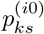 (*t*_*i*_) and 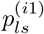 (*t*_*i*_) *∀i ∀k, l, s ∈ {*1, …, *d}*) can be performed independently, in parallel, and avoiding underflow issues for boundary conditions.

Overall, the PDF reconditioning technique can be applied to any model of the MTBD family, and facilitates its parameter estimation by separating master equation resolution (non-recursive and parallelizable) from likelihood calculation (recursive, but negligible in time cost compared to equation resolution). Recursive likelihood calculation can be performed with a standard pruning algorithm and rescaling techniques to control for potential underflow. For parameter estimation on trees with known node states (e.g., from metadata, or because they were generated by an MTBD process in which only one state can transmit or get sampled, like the BDEI model), tree likelihood can be calculated with a non-recursive formula in a logarithmic form (Eq. (8)), fully avoiding underflow.

### 2.1 Efficient parameter and CI estimation for the BDEI model

We applied our theoretical findings to implement a fast and efficient parameter estimator for the BDEI model (which we called PyBDEI). It estimates the BDEI model parameters Θ = (*μ, λ, ψ, ρ*) ∈ ℝ^4^ for a forest *ℱ* comprising *f ≥*1 observed trees in the maximum-likelihood framework, where one of the parameters in Θ is fixed (for identifiability reasons). The number of hidden trees *u* can be either given by the user, or estimated from BDEI model parameters (as in Eq. (6)).

Once the optimal parameter values are found, we calculate their confidence intervals (CIs) using Wilks’ method (Wilks, 1938). For each non-fixed parameter *p ∈* Θ, we calculate its 95%-CI as including the values 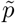 such that 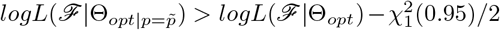, where 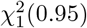 is the value of chi-squared distribution with 1 degree of freedom corresponding to the significance level of 0.95 (i.e., 3.84). 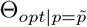 corresponds to the maximum-likelihood value for the other non-fixed parameters when 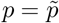.

### 2.2 Performance on simulated data and comparison to other tools

To assess the performance of our maximum-likelihood estimator PyBDEI, we used the simulated data from (Voznica et al., 2022), where we generated 100 medium trees with 200–500 tips under the BDEI model, with the parameter values sampled uniformly at random within the following boundaries: incubation period 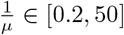, 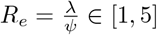, infectious time 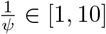, sampling probability *ρ ∈* [0.01, 1[. These trees were evaluated with the standard Bayesian method BEAST2 (Bouckaert et al., 2019) and the deep learning-based estimator PhyloDeep (detailed configurations are described in Materials and Methods). Additionally 100 large trees (5 000–10 000 tips) were generated for the same parameter values, and assessed with PhyloDeep in (Voznica et al., 2022). PhyloDeep’s maximal pre-trained tree size is 500 tips, however for larger trees it estimates BDEI parameters by (i) extracting the largest non-intersecting set of subtrees of sizes covered by the pre-trained set (50–500 tips), (ii) estimating parameters on each of the subtrees independently, and (iii) averaging each parameter’s estimate over the subtrees (weighted by subtree sizes).

To evaluate PyBDEI performance on forests, we additionally generated two types of forests for the large data set. The first type of forests was produced by cutting the oldest (i.e., closest to the root) 25% of each full tree, and keeping the forest of bottom-75% subtrees (in terms of time). We hence obtained 100 forests representing sup-epidemics that all started at the same time (*t* = 0, as in Fig. 3a). These forests contained 1 *−* 114 observed trees each, with a total of 5 031 *−* 9 953 tips.

The second type of forests represented epidemics that started with multiple introductions happening at different times (as in Fig. 3b). To generate them we (1) took the parameter values Θ_*i*_ corresponding to each tree *𝒥*_*i*_ in the large dataset (*i ∈ {*1, …, 100*}*), (2) calculated the time *T*_*i*_ between the start of the tree *𝒥*_*i*_ and the time of its last sampled tip, (3) kept (3.1) uniformly drawing a time *T*_*i,j*_ *∈* [0, *T*_*i*_] and (3.2) generating a (potentially hidden) tree *𝒥*_*i,j*_ under parameters Θ_*i*_ till reaching the time *T*_*i,j*_. Steps (3.1) and (3.2) were repeated till the total number of sampled tips in the generated trees reached at least 5 000: 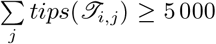. The resulting forest *ℱ*_*i*_ included those of the trees *𝒥*_*i,j*_ that contained at least one sampled tip (i.e., observed trees). These forests contained 1 −4 582 observed trees each, with a total of 5 000 −9 897 tips, and 0 −10 687 hidden trees.

We applied PyBDEI to these data sets, and compared the results to those reported for BEAST2 and PhyloDeep in (Voznica et al., 2022). For the large data set, we applied PyBDEI to full trees, but also to the two types of forests.

We calculated the relative error (normalized distance between the estimated and the target values: 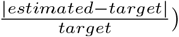 and the relative bias 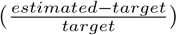 for each parameter on each tree/forest. Average relative errors for PyBDEI were ≤ 13% on the medium trees and *≤* 2% on the large trees (hence decreasing with the data set size, as expected), and well centered around zero (i.e., unbiased), as shown in Figure 4. The relative 95%-CI width 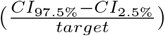 also decreased: from *∼* 0.5 on the medium data set to *∼* 0.1 on the large one. The target values of rates *μ, λ* and *ψ* were within the estimated CIs in correspondingly 92%, 89%, and 98% of cases on the medium data set, and in 95%, 90% and 95% of cases on the large one.

**Figure 4.**
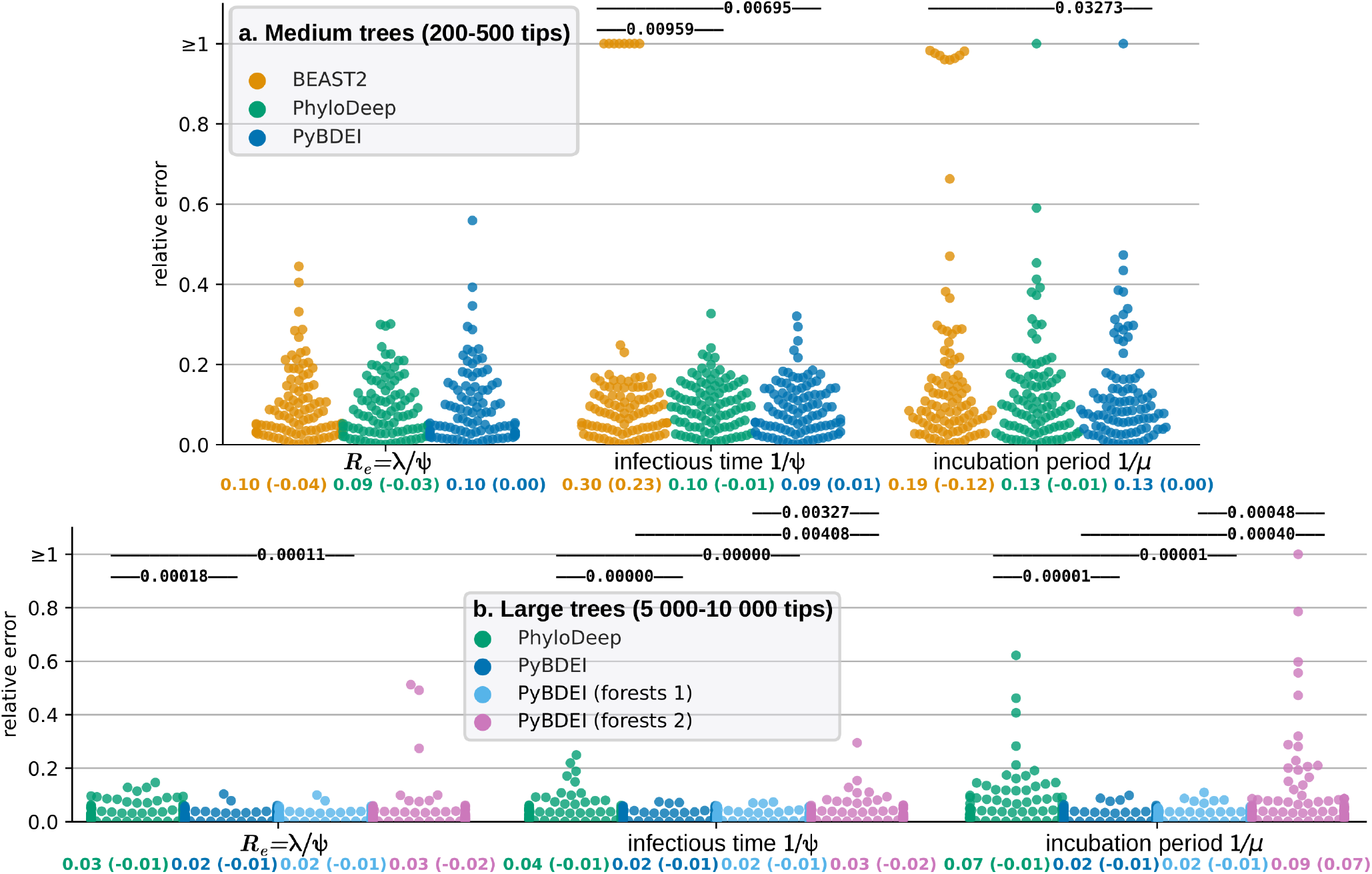
Comparison of inference accuracy of different methods on the medium (200-500 tips, top) and large (5 000–10 000 tips, bottom) 100-tree data sets. For the medium data set BEAST2 (in orange), PhyloDeep (in green) and our estimator (PyBDEI, in blue) are compared. Two (out of 100) trees of the medium data set on which BEAST2 did not converge after 10^6^ MCMC steps are excluded from the analysis and not shown in the figure. For large trees (*>* 500 tips), PhyloDeep extracts the largest non-intersecting set of subtrees of 50–500 tips, estimates parameters on each of the subtrees independently, and averages each parameter’s estimate over the subtrees (weighted by subtree sizes). We assessed our method on full trees (dark-blue), on forests obtained from the full trees by removing the oldest (closest to the root) 25% (in terms of height) of those trees (forests 1, light-blue), and on forests whose trees were generated using varying sampling period durations (forests 2, pink). We show the swarmplots (colored by method) of relative errors for each test tree/forest and parameter, which are measured as the normalized distance between the median a posteriori estimate by BEAST2 or a point estimate by PhyloDeep/PyBDEI and the real value. Average relative error (and in parentheses average relative bias) are displayed for each parameter and method below their swarmplot. The accuracy of the methods is compared by a paired z-test; *p <* 0.05 are shown above each method pair; non-significant p-values are not shown.

To assess method accuracy we calculated p-values based on two-sided z-test for each parameter and method pair. On the medium data set for *R*_*e*_ all the methods performed in a comparable way. For the infectious time, 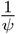, PyBDEI was at least as accurate as PhyloDeep and more accurate than BEAST2 (p-value *<* 0.05). For the incubation period, 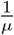, PyBDEI was more accurate than both other methods (see Fig. 4). On the large data set BEAST2 was inapplicable due to computation times (57 CPU hours were already required for each medium tree, on average), while PyBDEI was more accurate than PhyloDeep, both using full trees and the forests of the first type (i.e., where all the trees started at the same time, p-value *<* 0.01 for all the parameters, see Fig. 4). On the forests of the second type (i.e., trees starting at different times) PyBDEI’s performance was comparable to the one of PhyloDeep for all the parameters. While the mean relative errors were low (*<* 10%), PyBDEI performed worse (p-value *<* 0.01) on forests of type 2 than on full trees or forests of type 1 for the infectious period and incubation time. This can be explained by the fact that the starting times of the hidden trees in forests of type 2 were not known and hence needed to be approximated. Moreover, forests of type 2 contained less branches (mean 13 155) than forests of type 1 (14 896) or full trees (14 969), and hence less data for parameter inference.

For two trees in the medium data set, BEAST2 did not converge after 10^6^ Markov Chain Monte Carlo (MCMC) steps: We did not include these two data points in the analysis. PyBDEI performed well on these two trees: real parameters were withing estimated confidence intervals, relative errors for *R*_*e*_ *<* 15%, relative errors for incubation period and infectious time *<* 10%.

There were also several trees where BEAST2 seems to have converged to a local optimum (estimates with relative errors close to 1). To investigate this hypothesis further, we calculated the tree likelihoods for the real parameter values and those estimated by the three methods (Table 1). Indeed, BEAST2 had a likelihood lower than the one obtained on real values for 8% of trees in the medium dataset (8 out of 98 trees on which BEAST2 converged), which corresponds to a local optimum. These 8 data points correspond to the high BEAST2 relative error values (≥1 for infectious time and *>* 0.9 for incubation period) shown in Fig. 4. PyBDEI estimates had a higher or equal likelihood than any other method and real values for all the trees of both data sets, suggesting that PyBDEI reaches the global optimum of the likelihood function. On the medium dataset, PyBDEI estimates had an equal likelihood to the ones of BEAST2 for 85% of trees, and a higher likelihood for the other 15% of trees. Comparing to PhyloDeep, PyBDEI estimates had an equal likelihood for 48% of trees, and a higher likelihood for the other 52% of trees. Interestingly, PhyloDeep, while being a likelihood-free method, performed really well on the medium data set: on 81% of trees it estimated parameters with higher or equal likelihood to the one of the real parameters. On the large data set it performed worse in terms of likelihood, estimating parameters with higher or equal likelihood to the one of the real parameters only on 29% of trees. It could however be explained by the fact that PhyloDeep estimated parameters on each of the smaller (50-500 tip) subtrees selected by its subtree picker procedure, and averaged the result, instead of being retrained on large 5 000-10 000-tip trees.

**Table 1:**
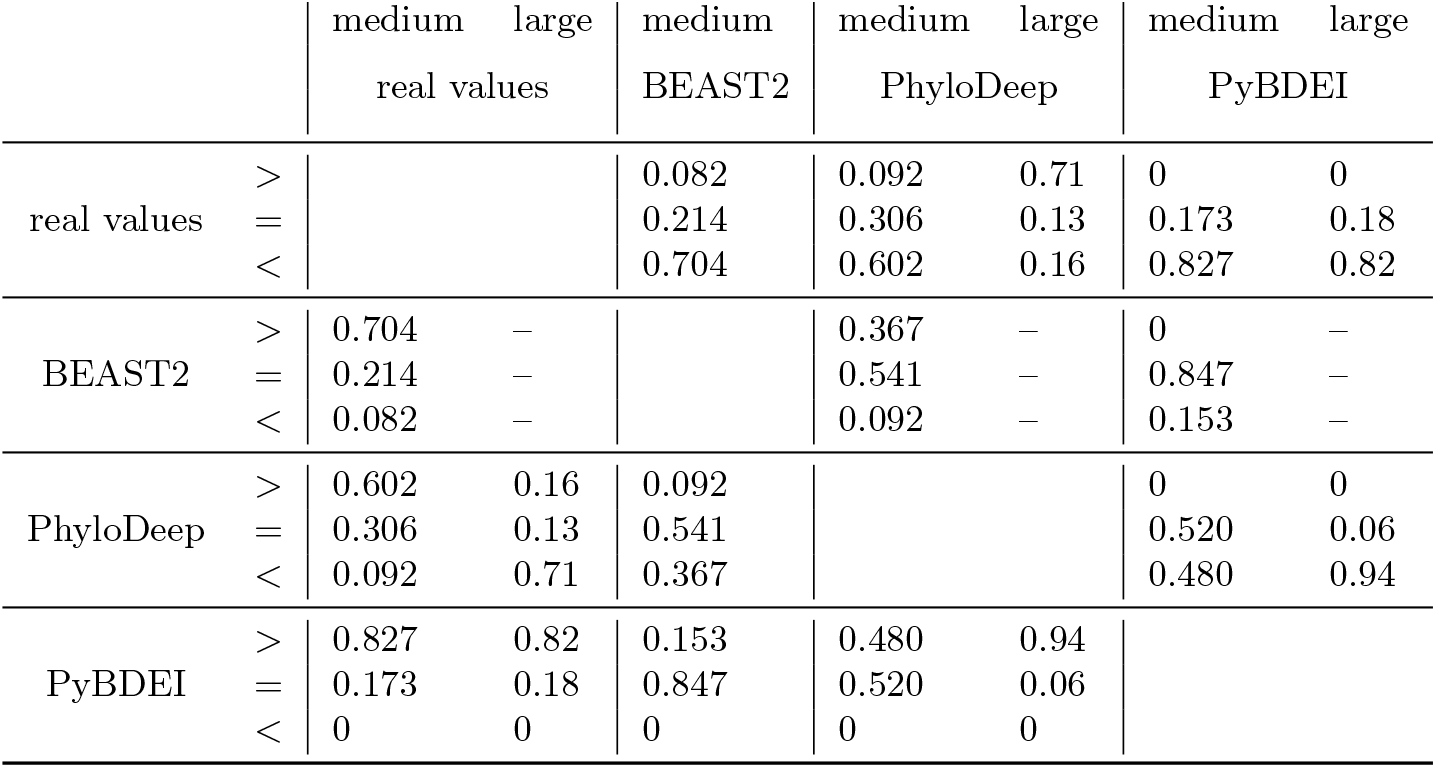
Tree likelihood comparison between the real parameter values and those estimated by different methods on medium and large data sets. The value provided in row i, column j indicates the proportion of trees for which the likelihood with parameters estimated by method i is either higher (sub-row *>*), equal (sub-row =) or greater (sub-row *<*) than the likelihood estimated by method j. For example, 0.847 in the row “BEAST2 =“ and the column “PyBDEI medium” means that the estimates of BEAST2 had an equal likelihood to those of PyBDEI on 84.7% of trees of the medium data set. According to a sign test, the differences between all the pairs of methods are significant (*p − value <* 0.01).

In terms of time, on the medium data set PyBDEI needed on average 4 seconds per tree on 1 CPU, and converged in 864 iterations (including CI calculation). These times cannot be directly compared to BEAST2 times, as BEAST2 performs a Markov Chain Monte Carlo (MCMC) parameter space exploration instead of looking for the optimum (as PyBDEI does), hence requires many more steps: for 10^6^ MCMC steps it took on average 57 CPU hours. While the number of MCMC steps could probably be reduced for some runs, for two out of 100 trees it was not sufficient for convergence. Implementing likelihood calculation with our new MTBD formulation and targeted numerical analysis methods, could be helpful in Bayesian context as well: comparing time per iteration (which is roughly time per likelihood calculation), our optimizer required ∼0.01 CPU seconds, while BEAST2 took one order of magnitude longer: ∼0.2 CPU seconds. This is probably due to the fact that BEAST2 uses the general MTBD model formulation, configured for BDEI, while our implementation uses a BDEI-tailored implementation. Moreover, the ODE reconditioning allows to avoid underflow errors (and rescaling efforts during tree pruning (Scire et al., 2022)), which could also play a role. Using several CPUs would allow for an even larger gain (see the results on the large data set below). PhyloDeep took 0.2 CPU seconds per tree, which is faster than our method’s time but does not include the training time of deep learning predictors (hundreds of hours). To our knowledge, the only other available maximum-likelihood estimator for BDEI is implemented in the TreePar package (Stadler and Bonhoeffer, 2013). However, as it suffers from underflow issues for BDEI already on trees of small size, its developers suggest using BEAST2 instead (private communication).

The average time of PyBDEI convergence on the large data set was 2 minutes 28 seconds minutes on 1 CPU, and required 960 iterations. Parallelization on 2 CPUs reduced it to 1 minute and 24 seconds (1.8 times faster). The speed up is close to the number of cores, which shows the efficiency of parallelization of master equation resolution despite the pre- and post-processing steps (tree reading, distribution of jobs between the threads, combining their results), which are always performed on one CPU. This suggests that our estimator will be easily applicable to much larger trees.

As the BDEI model requires one of the parameters to be fixed in order to become asymptomatically identifiable (Stadler, 2009), we fixed *ρ* to the real value, both in (Voznica et al., 2022) and in the comparison described above. However, to assess PyBDEI performance with other parameters fixed, we estimated parameters for trees in the large data set under three additional settings: with (1) *μ*, (2) *λ*, or (3) *ψ* fixed to its real value. The results are shown in Fig. S1. Average relative errors were ≤4% for all parameters when *μ* was fixed to the real value, ≤3% when *λ* was fixed, ≤2% when *ψ* or *ρ* were fixed. For estimates of *ρ* we calculated absolute errors |*estimated −target*| instead of relative ones: their average was 0.02 for fixed *μ* or *λ*, and 0.01 for fixed *ψ*. Hence, the estimations can be successfully performed with any of the parameters being fixed, but fixing *ψ* or *ρ* might be particularly useful. Moreover, these parameters are relatively easy to estimate with real data (e.g., patient observations for *ψ*, and proportion of sampled cases among the declared ones for *ρ*).

Finally, we assessed the impact of the number of hidden trees *u* on the parameter estimation. We estimated parameters with *u* = 0, and *u* being estimated from model parameters on the two types of forests of the large dataset. For forests of type 2 (where trees started at different times), we additionally compared *u* estimations using minimum, mean, median and maximum of observed tree-specific times (see Eq. (5)). Note that the minimum time and *u* = 0 represent the two extremes, as the probability of a tree to stay fully unobserved decreases with time. The relative errors for different parameters are shown in Fig. S2. While estimating *u* using mean, median or maximum times seems to have a slightly smaller relative error for *R*_0_ (3% vs. 4% for minimum time or *u* = 0), these differences are non-significant. Overall, *u* seems to have little impact on parameter estimation.

The assessment of our maximum-likelihood estimator on simulated data shows that it opens new possibilities to fast and accurate analyses of extremely large data sets, while being flexible with respect to parameter settings (e.g., the parameter to be fixed). The use of forest make it possible to focus on specific part of a large tree, e.g., the most recent period, the subtrees corresponding to a given region or country, or on the origin of the epidemics.

## 3 Application: Ebola in Sierra-Leone

Using PyBDEI, we analysed the 2014 Ebola epidemic in Sierra-Leone (SLE). Ebola virus features an incubation period (reported by the World health Organisation (WHO) to take between 2 and 21 days (WHO, 2021)). Using statistical methods based on time series of reported Ebola cases, the incubation period of Ebola during the 2014 SLE epidemic was previously estimated to be around 10–11 days, the infectious time around 4–5 days, and the reproduction number to decrease from around 2 in the beginning of the epidemic to values close to 1 by late 2014 (due to control measures) (Team, 2014, 2015; Rivers et al., 2014) (see also (Van Kerkhove et al., 2015) for a review).

Sequence data could improve and complement these estimates. However, the existing phylodynamic study of these parameters was limited by the data set size: Stadler and Bonhoeffer (2013) applied the BDEI model to the early spread of Ebola in SLE by analysing 72 Ebola samples from late May to mid June 2014 (sequences from Gire et al. (2014)). They estimated the expected length of the incubation period to be 4.9 days (median; 95% HPD 2.1-23.2), and the infectious time of 2.6 days (median; 95% HPD 1.2-7).

To show the power of phylodynamic analyses on larger data sets, we took the 1 610-sequence alignment and metadata (sampling times and countries) that were used in the study by Dudas et al. (2017), who analysed the factors that spread the 2014–2016 Ebola epidemic in West Africa. Using these data, we reconstructed a time-tree of the Ebola epidemic in West Africa, which we then used to extract a forest of time-subtrees representing the Ebola epidemic in SLE between July 30, 2014 (when the SLE government began to deploy troops to enforce quarantines (News24, 2014)) and September 7, 2015 (the last SLE sample in the data set). This was done to obtain a forest of subtrees with a homogenous health policy (after July 30). The details on the forest reconstruction are given in Materials and Methods. To check for robustness of the estimates, forest reconstruction was performed 10 times, obtaining slightly different forests.

We estimated the BDEI parameters on these 10 forests. As the BDEI model requires one of the parameters to be fixed for identifiability (Stadler, 2009), we performed the estimations fixing the sampling probability *ρ*. We estimated *ρ* as the proportion of cases represented by our forests (853–854) with respect to the total number of SLE Ebola cases reported by the Centers for Disease Control and Prevention (CDC) (CDC, 2020) between September 8, 2015 (the closest date to the last SLE sample in our data set, 13 683 cases) and July 31 (the day following the quarantine measures start, 533 cases): *ρ ≈*854*/*(13 683 −533) ≈0.065 (calculated independently for each forest). To check the robustness of the predictions with respect to this estimation of *ρ*, we additionally estimated the parameter values assuming 20% more (15 780, *ρ ≈*0.054) and 20% less (10 520, *ρ ≈*0.081) total cases. For each of these settings we performed three estimations: (1) with the number of unobserved trees being estimated, (2) with it being fixed (via setting the parameter *u*) to the difference between the total number of SLE Ebola cases reported by the CDC on July 31 (533) and the number of trees in the corresponding forest (varying between 143 and 174), and (3) with it being fixed to zero.

The results for different *ρ* values, estimated vs. fixed *u*, and trees were compatible, with intersecting CIs (see Table S1). We estimated the *R*_*e*_ value between 0.95 and 1, suggesting a contained epidemic (which is in a good agreement with the quarantine measures and the end of the epidemic in early 2016). The incubation period was estimated between 11 and 14 days. These estimates are fully compatible with the previous studies and allow to narrow down the WHO incubation period estimate (2–21 days, non-specific to the SLE epidemic). We estimated a very short infectious period 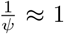 hour. While it does not correspond to the epidemiological estimates (4–5 days) reported in previous studies, it makes sense in the setting we are looking at. The BDEI infectious period corresponds to the time interval between the moment when a person becomes infectious and the moment when they cannot transmit anymore. In the beginning of an epidemic impossibility to transmit is typically defined by biological factors, such as healing or death, and corresponds to the epidemiological definition. However, it could also be influenced by logistic reasons, such as self-isolation. As we are looking at the lock-down period, with strict surveillance, it seems likely that a person who develops symptoms (i.e., passes from the exposed to the infectious state) is immediately detected and isolated, hence having very limited time to transmit on average. Our estimate therefore corresponds to this logistic scenario.

Note, that comparing the setting with *u* being fixed according to the CDC-declared case count to the one when it is estimated, *R*_*e*_ estimates were slightly smaller with *u* fixed (while the CIs intersected). As the number of cases is defined by the epidemic preceding the studied period, *u* might not correspond to the number estimated from the studied period parameters, and it might be more accurate to fix it.

Overall, the analysis took ∼1 hour for the reconstruction of forests and ∼1 minute per forest for the BDEI parameter estimation.

This application shows the advantages of PyBDEI not only in terms of calculation times, but also in terms of flexibility of input settings (extracting information from multiple trees).

## 4 Discussion

We proposed a highly parallelizable formulation of master equations for MTBD models. We also proposed an extension of the MTBD models to forests, to tackle situations where health policies change over time (providing a flexible alternative to the Bayesian skyline), as well as situations of multiple (not necessarily simultaneous) pathogen introductions to a country of interest. The extension to forests does not introduce additional model parameters, however, when available, it allows to incorporate external data on the number of infectious cases at the start of the forest.

The peculiar properties of original MTBD equations permitted us to rewrite them in a branch-specific way. This representation features simple boundary conditions with 0 or 1 values, and avoids numerical and underflow problems that could occur in the original system due to very small positive values of the boundary conditions. Even more importantly, our branch-specific representation removes the recursive dependency between the equations corresponding to parent and child tree nodes, and permits their time-consuming resolution to be performed in parallel and independently. The results can then be combined into tree likelihood with a nearly standard pruning algorithm. While the likelihood-combining step in the most general MTBD case remains recursive, its time cost should be negligible in comparison with the recursive master equation resolution used in previous studies. Moreover, for cases where tree node states are known we obtained an explicit likelihood formula, which can be represented in a logarithmic form (Eq. 8) to avoid potential underflow issues during the likelihood-combining step. Tree node states could be known from metadata, or because they were generated by an MTBD process in which only one state can transmit or get sampled.

We implemented our theoretical findings in a maximum likelihood parameter and CI estimator for the BDEI model, which is a special case of MTBD models, and one of the most useful models in epidemiology, being applicable to Ebola, Sars-CoV 1 and 2, Tuberculosis and other pathogens that feature an incubation period. Under this model tree node states are known, as only the infectious individuals can transmit their pathogen or get detected (after symptom onset). Our parameter estimator, PyBDEI, drastically increases parameter optimization performance, accuracy and speed with respect to previously available estimators.

We applied our estimator to the 2014 Ebola epidemic in Sierra Leone, after the introduction of quarantine measures. The analysis took *<* 2 hours (the majority of which was the tree reconstruction). The obtained estimates of epidemiological parameters are in agreement with what we now know about this epidemic. In particular, the estimate of *R*_*e*_, slightly below 1, suggests a contained epidemic, and indeed Sierra Leone was declared Ebola-free in early 2016, a few months after the sampling date (September 7, 2015) of the most recent Ebola sequence in our data set.

The accuracy of estimations improves with the data set size (as expected, see our simulations). In the world of rapidly growing sequencing data sets (Hodcroft et al., 2021), we can gain important insights on epidemic spreads by harvesting all available information. PyBDEI is applicable to very large data sets (2 minutes on a 10 000-tip tree), making parameter and CI estimation instantaneous with respect to phylogenetic tree reconstruction times (hours or even days). Our approach could be easily used in a Bayesian setting as well, and could potentially be implemented in BEAST2.

As the MTBD models are epidemiological analogues of the ClaSSE-like models, our findings could also be easily transferred to the macroevolution domain. Our parallelizable MTBD model formulation is closely related to general matrix-based flow framework recently proposed by Louca and Pennell (2020). Using this framework, they implemented an efficient parameter estimator *castor* for MuSSE-like models, however this type of models does not cover cladogenetic changes possible in ClaSSE-like and MTBD models. Moreover, our approach, thanks to forests, allows for multiple introductions, for example, of an epidemic in a given country, or of species from the same clade within a given ecological realm), which could be useful in the macroevolution domain. With rapidly growing genome sequence data, castor and PyBDEI open way to fast and accurate parameter estimations for ecology and epidemiology.

## 5 Materials and Methods

**Reconditioned BDEI master equations**

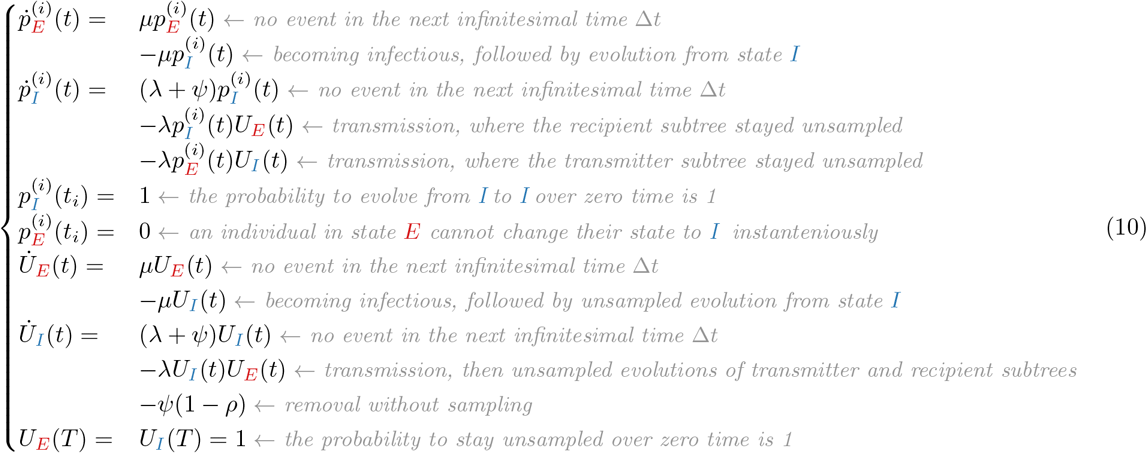

### 5.1 Equivalence between Equations (2) and (8)

The likelihood Equation (2) for a tree *𝒥* is recursive, and when using 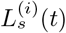 (*s* ∈ {1, …, *d}*) needs to be resolved with a pruning algorithm while climbing the tree. However, for a tree with known node states, we can transform it into a non-recursive Equation (8) with 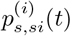, by alternating *replacement* and *unfolding* steps. A replacement step consists in replacing 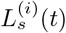 with 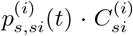 and is followed by an unfolding step. An unfolding step either (if *i* is a tip) unfolds 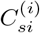 into *ψ*_*si*_*ρ*_*si*_ and stops; or (if *i* is an internal node) unfolds 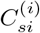 into 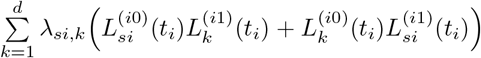 and proceeds with replacements. In Equation (11) we show the transformation process.

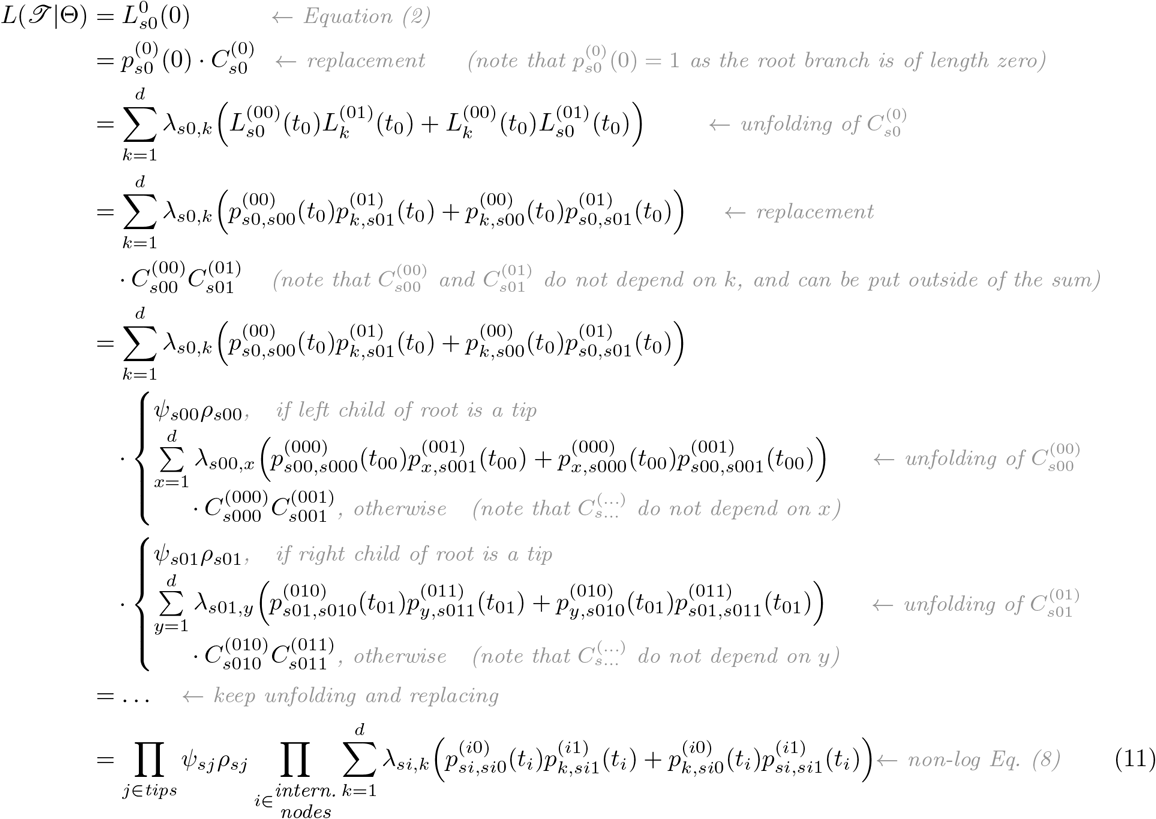

### Stationary distribution

Stationary state distribution Π = *{π*_*E*_, *π*_*I*_ *}* : *π*_*E*_ + *π*_*I*_ = 1 corresponds to the ratios of states *E* and *I* at a given time *t* after these ratios stopped changing (assuming that this may happen).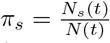, where *N*_*s*_(*t*) is the number of individuals of type *s ∈{E, I}* and *N* (*t*) is the total number of infected (infectious or not) individuals at time *t*. Hence, the derivative of the number of individuals of type *s* is proportional to the derivative of the total number of infected individuals:

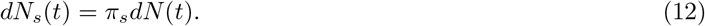

The number of individuals in state *I* increases due to becoming infectious of individuals in state *E* and decreases due to removal, while the number of individuals in state *E* decreases due to becoming infectious and increases due to transmissions:

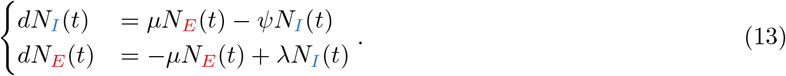

Becoming infectious changes the corresponding individual’s state but does not affect the total number of infected individuals, transmissions increase the total number, and removal decreases it. Note that only individuals in state *I* can transmit or be removed:

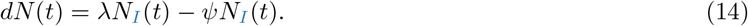

Combining [13] and [14] we rewrite [12] as a system of multivariate algebraic equations:

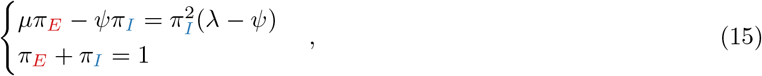

from which we derive a quadratic equation for 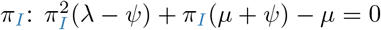, and the following stationary distribution:

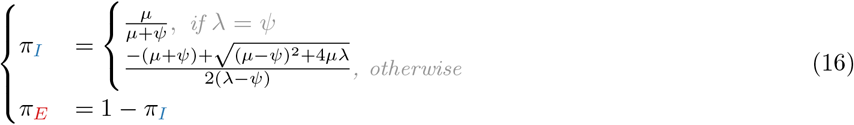

### PyBDEI, its code and data availability

Parameter estimation starts with a preprocessing step of reading the input trees, calculating the time *t*_*i*_ at each node *i* and memorising the association between each node and its child nodes: *i →* (*i*0, *i*1). This step requires one tree traversal, and its time is negligible with respect to the numerical master equation resolution performed in the next steps.

The estimation then proceeds with a search for the optimal parameter set 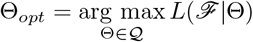, where 𝒬 is the set of admissible parameter values. We use the globally-convergent method of moving asymptotes (Svanberg, 2002) for the optimization. At each optimization step *k* the corresponding likelihood *L*(*ℱ*|Θ_*k*_) needs to be calculated, which implies calculating 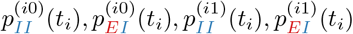 for each of the *n − f* observed internal nodes of *ℱ*, and combining them as in the forest likelihood formula (Eq. (4)). The reconditioned version of BDEI master equations (10) for the parameter values Θ_*k*_ can be resolved in parallel for each of the 2*n − f* observed forest nodes (differing in the times *t*_*i*_ of their boundary conditions).

To resolve these master equations numerically, we start by separating the self-defined subsystem for the unknowns *U*_*E*_(*t*) and *U*_*I*_ (*t*) from the rest of System (10), and calculate it independently. Taking into account the fact that the equations in System (10) are either linear (for 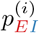 (*t*) and 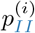 (*t*)) or quadratic (for *U*_*E*_(*t*) and *U*_*I*_ (*t*)) the use of an implicit scheme is simple. We chose implicit schemes with an automatic computation of the time step, such that the error is less than a given tolerance. In our implementation, we used the implicit Euler scheme (Butcher, 2016) for solving the linear equations and the Crank-Nicolson implicit scheme (Crank and Nicolson, 1947) for the non-linear ones. This permits us to avoid possible stability time restrictions and only choose the time steps for precision.

The core of our estimator is implemented in C++ and uses the NLopt library (Johnson) for non-linear optimization. The parallelization is achieved with the C++ `thread_pool` tools (Williams, 2012). To facilitate the use of our estimator in Python and perform additional validation of input trees, we wrapped the core estimator into a Python 3 library PyBDEI. PyBDEI uses ETE 3 framework for tree manipulation (Huerta-Cepas et al., 2016) and NumPy package for array operations (Harris et al., 2020).

Our estimator is available as a command-line program and a Python 3 library via PyPi (pypi.org/project/pybdei), and via Docker/Singularity (hub.docker.com/r/evolbioinfo/bdei). Its source code, the simulated and real data used for its assessment, as well as the Snakemake (Köster and Rahmann, 2012) data analysis pipelines, and the installation and usage documentation are available on GitHub at github.com/evolbioinfo/bdei. The simulated data and trees generated for Ebola epidemic are also available from the Dryad Digital Repository: https://doi.org/10.5061/dryad.r7sqv9sgx.

### BEAST2 and PhyloDeep settings

BEAST2 (v2.6.2 with package bdmm (Scire et al., 2022) v1.0) was configured for 10^6^ MCMC steps with the following priors: *μ ∈U* (0.02, 5.0), *ψ ∈U* (0.1, 1.0), and *ρ* fixed to the real value (∈ [0.01, 1[, different for different trees). The initial values in the MCMC were set to the medians used in the PhyloDeep training set, namely *μ* = 2.51, *ψ* = 0.55, and *R*_0_ = 3. The tree was fixed to the real tree. For each tree, the Effective Sample Size (ESS) on all parameters was evaluated, and the median of a posteriori values was reported, corresponding to all recorded steps (i.e., actual MCMC steps spaced by 1 000) past the 10% burn-in. The simulations for which BEAST2 did not converge after 10^6^ MCMC steps (2%) were discarded from the analyses.

PhyloDeep (v0.2.51) was run with *ρ* fixed to the real value and Convolutional Neural Networks trained on the Compact Bijective Ladderized Vector full tree representation (CNN-CBLV).

The visualisations of the analyses of simulated data were performed with the Python 3 library seaborn (v0.11.2) (Waskom, 2021; Hunter, 2007).

### Tree reconstruction for Ebola SLE epidemic analysis

We reconstructed a maximum-likelihood phylogeny of 1 610 tips for the Ebola samples from (Dudas et al., 2017) with RAxML-NG (v1.0.2, GTR+G4+FO+IO) (Kozlov et al., 2019), and rooted it based on sampling dates using LSD2 (v2.4.1) (To et al., 2016). As Ebola’s mutation rate is slower than its transmission rate, the initial phylogeny contained 242 polytomies (i.e., multiple transmissions, which happened faster than the virus acquired a mutation, hence making them undistinguishable in the phylogeny). The BDEI model, on the other hand, assumes a binary tree. We therefore resolved these polytomies randomly (10 times, to check for robustness of the estimates) using a coalescent approach.

We then dated each of the 10 trees with LSD2 (To et al., 2016) (v2.4.1: github.com/tothuhien/lsd2/tree/v.2.4.1, under strict molecular clock with outlier removal) using tip sampling dates, and reconstructed the ancestral characters for country with PastML (Ishikawa et al., 2019) (v1.9.40, MPPA+F81).

Lastly, we extracted 10 SLE forests from these trees to represent the Ebola epidemic in SLE between July 30, 2014 (when the SLE government began to deploy troops to enforce quarantines (News24, 2014)) and September 7, 2015 (the last SLE sample in our dataset) by (1) cutting each tree on July 30, 2014 to remove the more ancient part (with a different health policy); (2) among the July-31-on trees, picking those whose root’s predicted character state for country was SLE (light-green branches at the level of July 31, 2014 in Figure S3); (3) removing the non-SLE subtrees (indicated with other colors in Figure S3) from the selected July-31-on SLE trees to focus on the epidemic within the country, without further reintroductions.

The reconstruction took 1 hour for the phylogeny, 10 minutes for tree dating, and 1 minute for country ancestral character prediction.

## Data Availability

All data produced are available online at https://github.com/evolbioinfo/bdei

## 7 Acknowledgements

The authors thank Dr Jakub Voznica for valuable discussions.

## 8 Author Contributions

O.G. initiated the project and provided critical assessment of project progression; A.Z. conceived the new master equation representation, extension to forests, applications to Ebola and simulated data, and coordinated the project; F.H. and Y.M. conceived the numerical approach for fast and efficient parameter optimization; F.H. implemented the numerical approach; A.Z. wrote the manuscript with input from all the authors; all authors discussed the intermediate and final results.

## 9 Appendix

**Fig. S 1:**
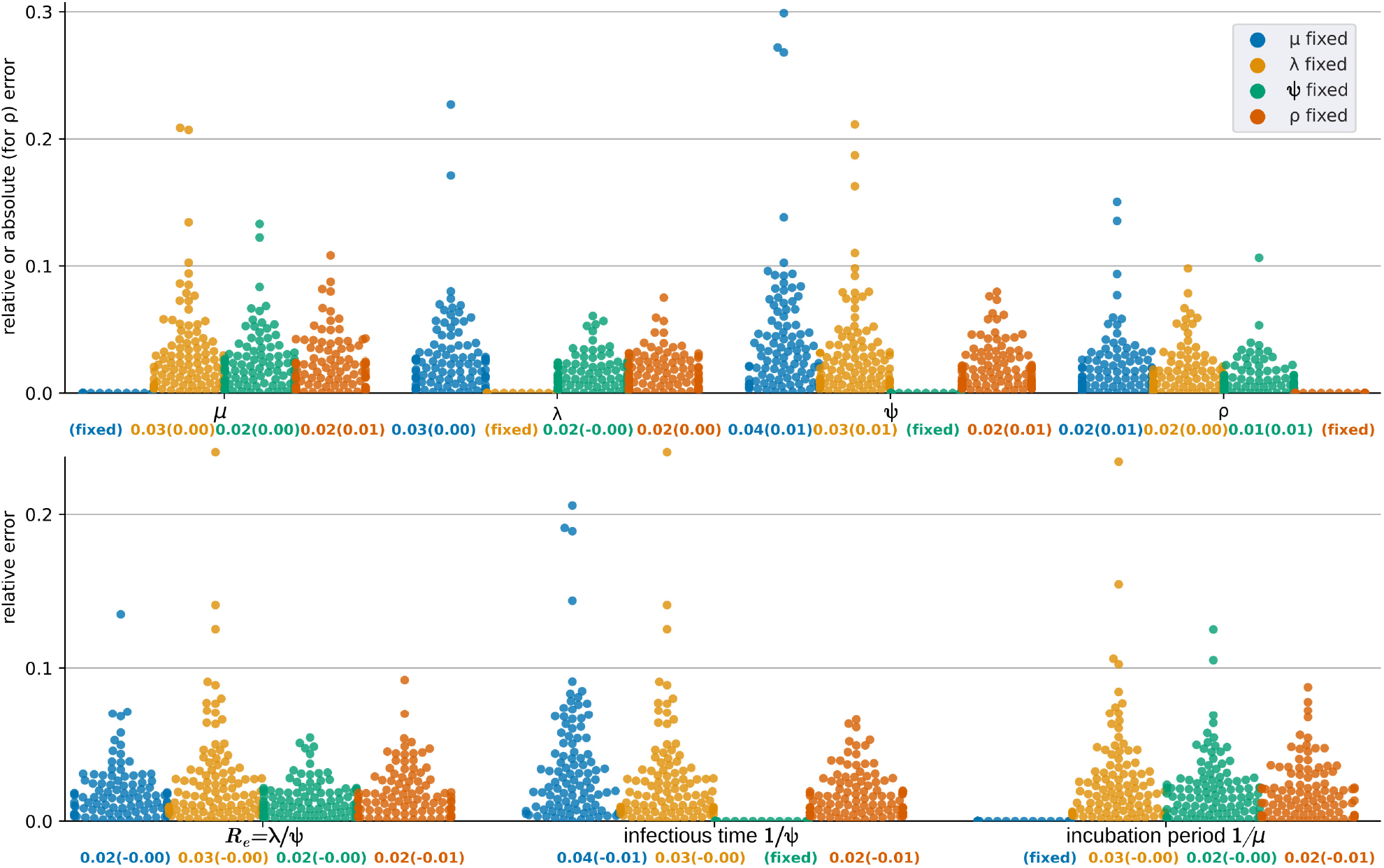
Comparison of inference accuracy on 100 test trees of 5 000–10 000 tips from (Voznica et al., 2022), with (blue) *μ* fixed to the real value, (yellow) *λ* fixed to the real value, (green) *ψ* fixed to the real value, (orange) *ρ* fixed to the real value. We show the swarmplots of relative errors for each test tree and parameter. Average relative error (and in parentheses average relative bias) are displayed for each parameter and setting below their swarmplot. For *ρ*, absolute errors and bias are shown instead of the relative ones.

**Table S 2:**
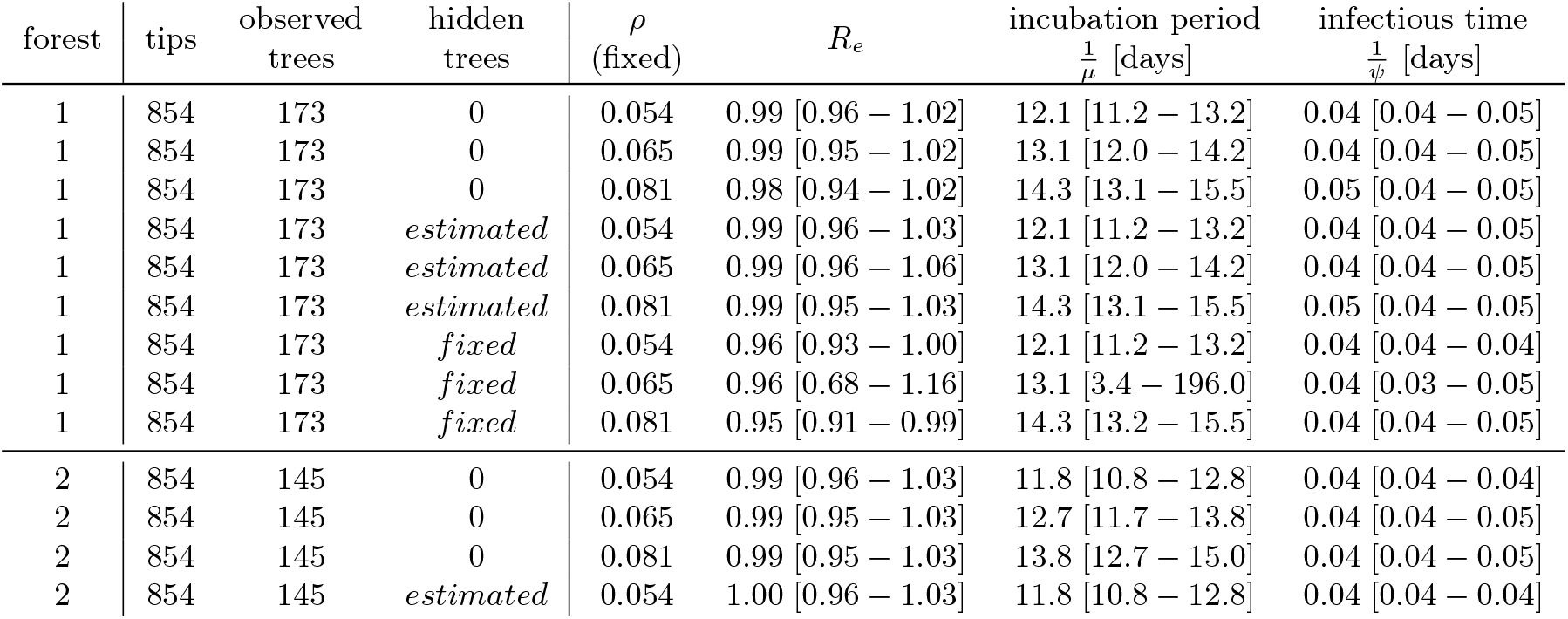
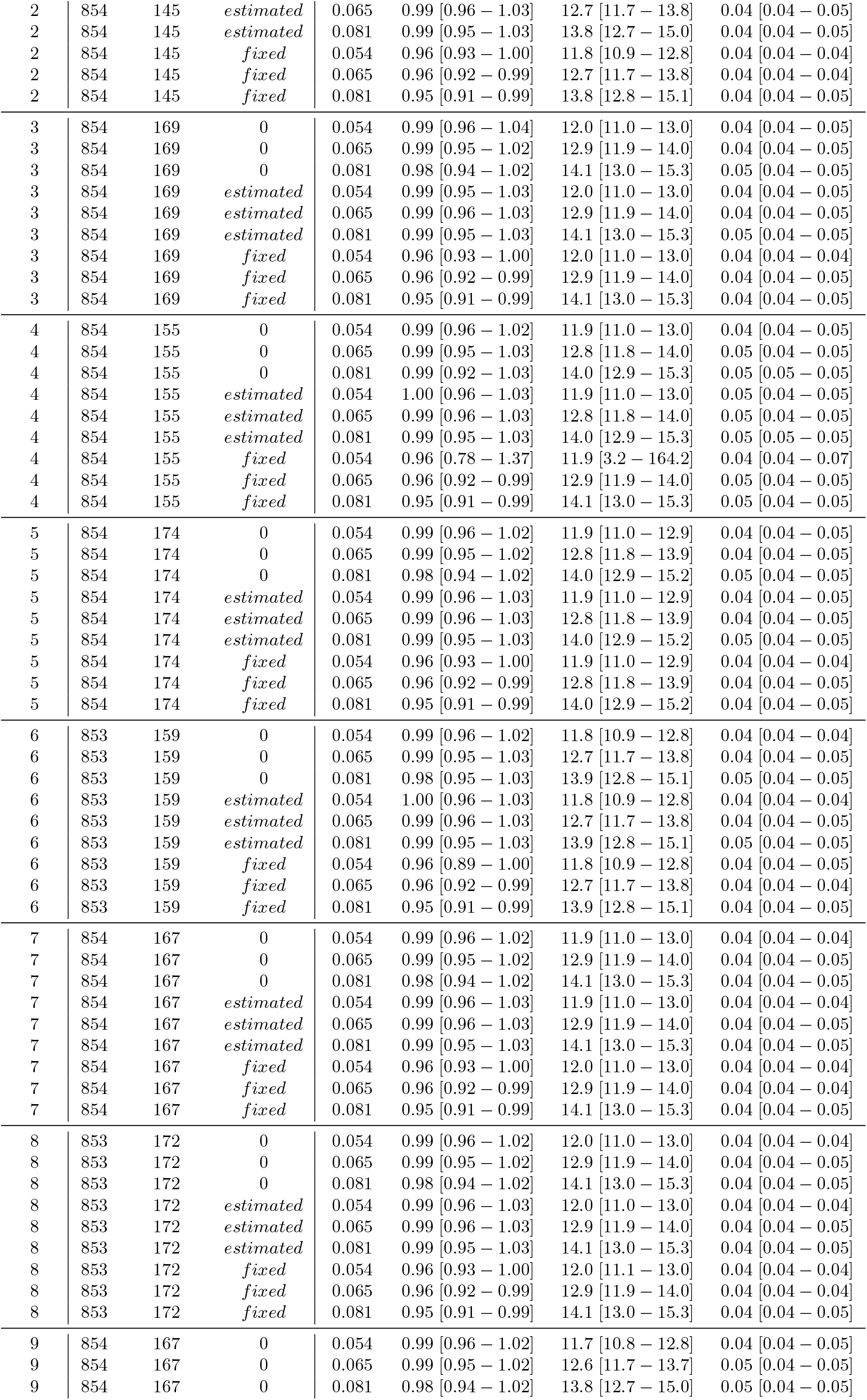
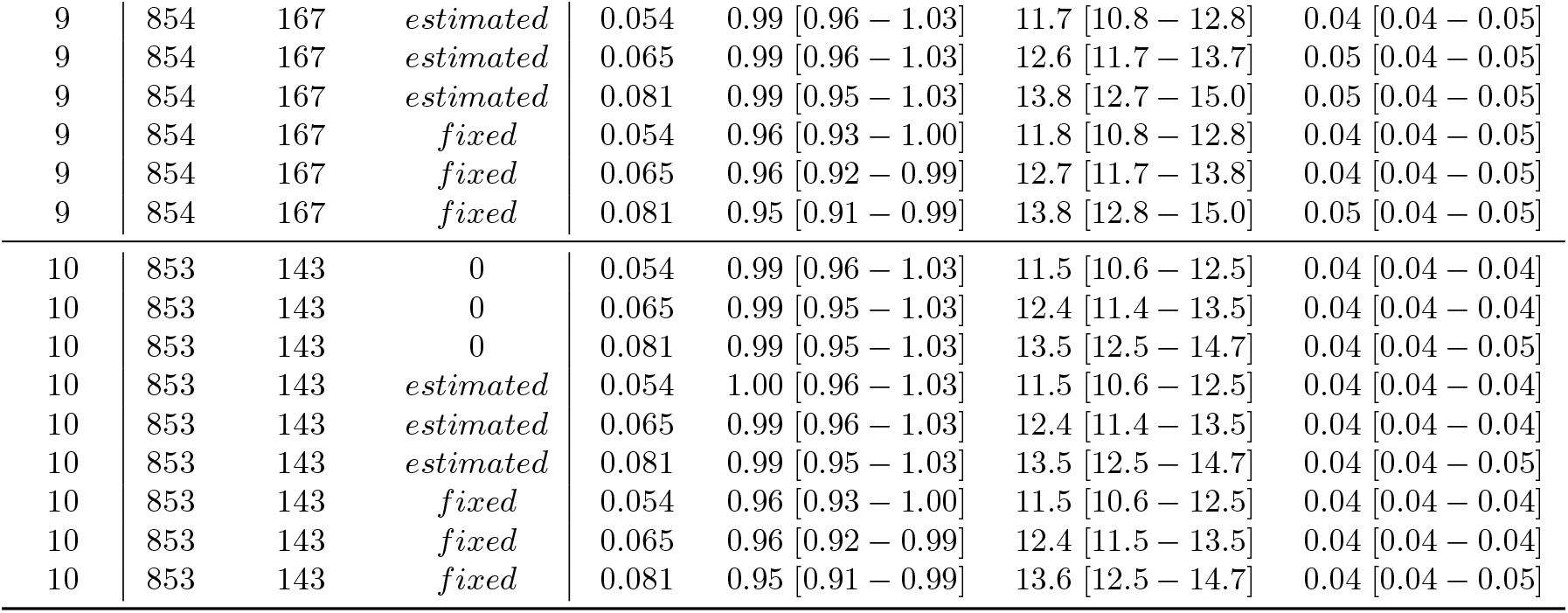
Parameters estimated for SLE 2014 Ebola epidemic.

**Fig. S 2:**
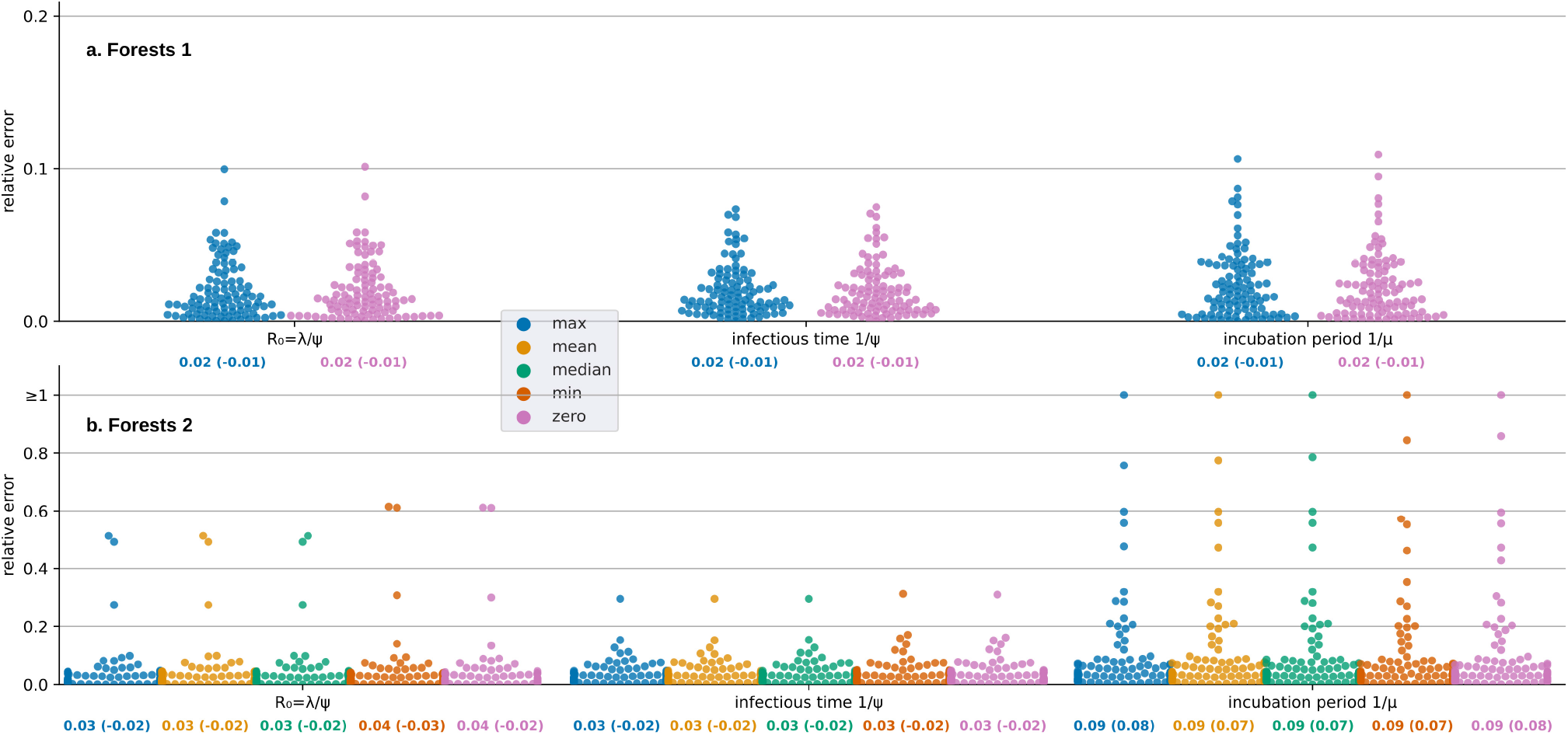
Comparison of inference accuracy on 100 forest of 5 000–10 000 tips of the large data set, with the number of hidden trees *u* fixed to zero (pink), or estimated using maximum (blue), mean (yellow), median (green) or minimum (orange) of observed tree times. The parameter *ρ* is fixed to the real value. We show the swarmplots of relative errors for each test forest and parameter. Average relative error (and in parentheses average relative bias) are displayed for each parameter and setting below their swarmplot. (a) Forests 1 represent forests whose trees started at the same time. (b) Forests 2 represent forests whose trees started at different times.

**Fig. S 3:**
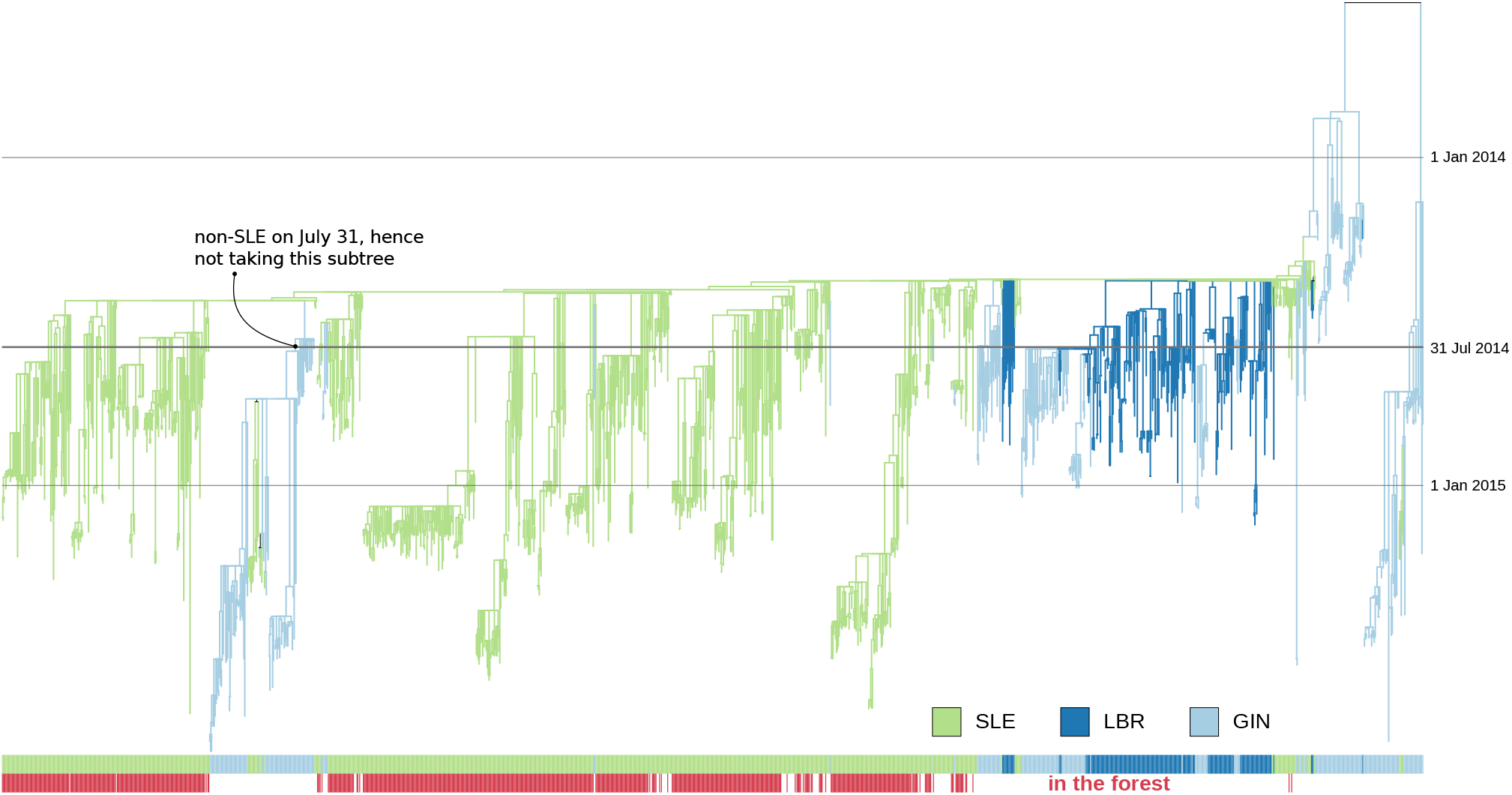
Ebola 2014 – 2016 epidemic time-scaled tree (data from (Dudas et al., 2017), polytomies are resolved as in forest 1) colored by country predicted by PastML (Ishikawa et al., 2019): Guinea (GIN) is light-blue, Liberia (LBR) is dark-blue, Sierra-Leone (SLE) is light-green. The bottom colorstrip shows the samples kept for the SLE 2014 epidemic analysis (forest 1): SLE samples that are directly related to the SLE epidemic of 31 July 2014 (start of quarantine) and sampled after this date. The SLE samples that were later reintroduced to SLE via other countries (e.g., in the indicated GIN-rooted subtree) were not included in the analysis.

